# The chronic wound virome: phage diversity and associations with wounds and healing outcomes

**DOI:** 10.1101/2022.01.05.22268807

**Authors:** Samuel Verbanic, John M. Deacon, Irene A. Chen

## Abstract

Two leading impediments to chronic wound healing are polymicrobial infection and biofilm formation. Recent studies have characterized the bacterial fraction of these microbiomes and have begun to elucidate compositional correlations to healing outcomes. However, the factors that drive compositional shifts are still being uncovered. The virome may play an important role in shaping bacterial community structure and function. Previous work on the skin virome determined that it was dominated by bacteriophages, viruses that infect bacteria. To characterize the virome, we enrolled 20 chronic wound patients presenting at an outpatient wound care clinic in a microbiome survey, collecting swab samples from healthy skin and chronic wounds before and after a single, sharp debridement procedure. We investigated the virome using a virus-like particle enrichment procedure, shotgun metagenomic sequencing, and a k-mer-based, reference-dependent taxonomic classification method. Taxonomic composition, diversity, and associations to covariates are presented. We find that the wound virome is highly diverse, with many phages targeting known pathogens, and may influence bacterial community composition and functionality in ways that impact healing outcomes.

**Importance:** Chronic wounds are an increasing medical burden. These wounds are known to be rich in microbial content, including both bacteria and bacterial viruses (phages). The viruses may play an important role in shaping bacterial community structure and function. We analyzed the virome and bacterial composition of 20 patients with chronic wounds. The viruses found in wounds are highly diverse compared to normal skin, unlike the bacterial composition, where diversity is decreased. These data represent an initial look at this relatively understudied component of the chronic wound microbiome.

## Introduction

Chronic wounds (i.e., those that fail to exhibit reasonable healing progress within an expected time frame) are a growing source of morbidity and mortality worldwide [1-3]. While not always infected, chronic wounds are frequently colonized by polymicrobial communities. Characterization of these communities is important for understanding the microbial content of wounds and the potential influence of the wound microbiome on healing outcomes. Recently, several culture-independent studies have characterized the extensive microbial diversity of skin and wounds [4-25]. These studies have found that wound communities are primarily composed of *Staphylococcus spp*., *Pseudomonas spp*., *Corynebacterium spp*., *Streptococcus spp*., *Anaerococcus spp*., and *Enterococcus spp*., and numerous low-abundance taxa. However, significant interpatient variability in the composition of the wound microbiome exists, which cannot be explained by covariates like age, race, sex, or wound etiology [9, 11]. Still, some studies have indicated that community composition may be associated with healing outcomes. Temporal instability and the transition between several distinct community structures was associated with positive healing outcomes [4], and communities with high proportions of aerobes and facultative anaerobes were associated with poor healing outcomes [26].

While the bacterial content of chronic wounds has been the subject of substantial study, the wound virome has received less attention. Previous work has determined that the human virome is mostly composed of bacteriophages, viruses that infect bacteria [27, 28]. The ability of bacteriophages to infect, kill, and modulate bacterial host function has been well-described [29-31], and the wound virome may therefore represent a previously understudied explanatory variable for inter-patient variability, healing outcomes, community dynamics, and pathogenicity [28, 32-34]. Despite its potentially influential role in the bacterial microbiome, studies of the virome are often hindered by insufficient sequencing depth. Although viruses and phages are highly abundant in number (outnumbering bacterial cells approximately 10:1), their typically small genome sizes result in a small fractional abundance of viral DNA sequences compared to prokaryotic and eukaryotic DNA. Therefore, virus-like particle (VLP) enrichment is necessary to obtain substantial sequencing depth of the viral fraction.

The viral fraction of the healthy skin microbiome has been studied [35, 36], but to date, only one study has employed a VLP enrichment method [37]. In this study, 91% of putative viral contigs for dsDNA viruses could not be taxonomically classified; among those that could be classified, most belonged to the order Caudovirales and targeted *Staphylococcus spp*., *Corynebacterium spp*., *Streptococcus spp*., *Propionibacterium spp*., and *Pseudomonas spp*., and the most common virus infecting humans was papillomavirus. Virome composition and diversity was associated with the skin site (e.g., sebaceous or moist, occluded or exposed), and exhibited high intra-personal variance but less temporal variance at a given site. Despite progress in describing the human virome in general [27, 28, 38], no studies have used VLP-enrichment methods and metagenomics to characterize the wound virome. The phage content of chronic wounds is of particular interest, as long-term community dynamics, which may be influenced by phages, might affect healing outcomes [4, 26].

Here, we characterize the chronic wound viromes of 20 patients presenting to an outpatient wound care clinic. Swabs were collected from chronic wounds before and after a single, sharp debridement event, along with a skin sample from the contralateral limb. For a detailed description of the patient cohort, see [26]. We previously reported characterization of the bacterial communities by Illumina sequencing of the V1-V3 loops of the 16S rRNA genes [26], including the finding that facultative anaerobes, particularly Enterobacter, were significantly associated with non-healing wounds. In the present work, we characterize the viral fraction of these samples. Samples were fractionated to enrich for VLPs while retaining a separate bacterial fraction, as described in [39]. The VLP fraction was characterized by shotgun sequencing, with read classification and taxonomic abundance analysis performed with Kraken2 and Bracken, respectively, using a custom database containing the latest NCBI Viral RefSeq genomes and the Joint Genome Institute’s IMG/VR viral metagenome database [40-43]. Viromes were analyzed using ecological diversity metrics and differential abundance analyses. We report the composition of dsDNA viromes from the chronic wounds as well as the contralateral skin sites, and find that wounds harbor significantly more diverse viral communities than skin, with most viruses being bacteriophages. Additionally, we identify specific taxonomic associations with wounds compared to skin, and for healing outcomes (healed vs. unhealed wounds). This study thus reports an investigation of the previously uncharacterized chronic wound virome.

## Results

### Read processing and classification

High-throughput sequencing of DNA from virus-like particles (VLPs) isolated from skin and wound samples, along with negative controls, resulted in 635,166,925 total paired end reads (9,623,741 ± 5,091,161 per sample, on average). Reads were quality- and adapter-trimmed, length-filtered, and joined, resulting in 556,645,739 pre-processed reads (8,434,026 ± 4,541,820 per sample, on average), which equates to an overall read retention rate of 86.64% (87.49 ± 9.92% per sample, on average). An initial assessment of overall taxonomic composition against the full NCBI RefSeq database indicated that most sequences had human or bacterial origin, though viral read abundances were still substantial (Figure S1).

To better assess the viral content of the samples, reads were re-classified with Kraken2 against the NCBI Viral RefSeq database, followed by classification against JGI’s IMG/VR database, a large, public viral metagenome repository [42, 43]. Taxon abundance estimates were again calculated with Bracken. Using this method, viral read classification was 379,992 ± 581,996 viral reads per sample, on average. Two types of taxonomic classifications were assigned to each hit: a viral species designation as assigned by NCBI or IMG/VR, if available, and a viral ‘type’ designation, which denotes host association for prokaryotic viruses or common viral family name for eukaryotic viruses. For phages, the viral type designation is also referred to in the report below as the presumed host species. Additional information regarding the curation of taxonomic designations can be found in the Methods.

To identify likely contaminants, negative control samples were prepared and sequenced in parallel with the true samples. At the ‘type’ level, Escherichia phage and unclassified viruses were found to be potential contaminants (Figure S2a), possibly due in part to experiments carried out in adjacent lab spaces. In the negative controls, at the species level, ambiguous Escherichia phage and unclassified taxa were the most abundant, followed by known lab strains including Escherichia viruses Lambda, DE3, T7, T4, and M13 (Figure S2b). An initial decontamination procedure was implemented with the R package Decontam, which identifies taxa that are more prevalent in negative controls than true samples [44]. Using the four sequenced negative control samples and a stringent threshold for declaring a contaminant, Decontam identified 39 potential contaminants. In addition, taxa corresponding to known lab strains were manually removed (Figure S3-4).

After decontamination, most of the virome could be assigned as a species or to a host organism. Nevertheless, many taxa had no known host association, accounting for 42.27±18.72% average relative abundance (Figure S5). Unless otherwise stated, the following results focus on the ‘defined’ fraction of the virome, which has viral species designation and/or ‘type’-level assignment of host species.

### Abundant viral species and phage hosts in skin and wound viromes

Skin samples exhibited high relative abundance of phages presumed to infect *Chryseobacterium, Neisseria, Staphylococcus, Yersinia, Bacillus, Pseudomonas, Salmonella, Corynebacterium*, and *Streptococcus* (Figure 1). Accordingly, the most abundant identifiable viral species on skin were unnamed *Neisseria, Yersinia, Bacillus, Corynebacterium, Streptococcus* and *Pseudomonas* phages (Figure S6). Among viruses that infect humans, papillomavirus was the most common.

**Figure 1:**
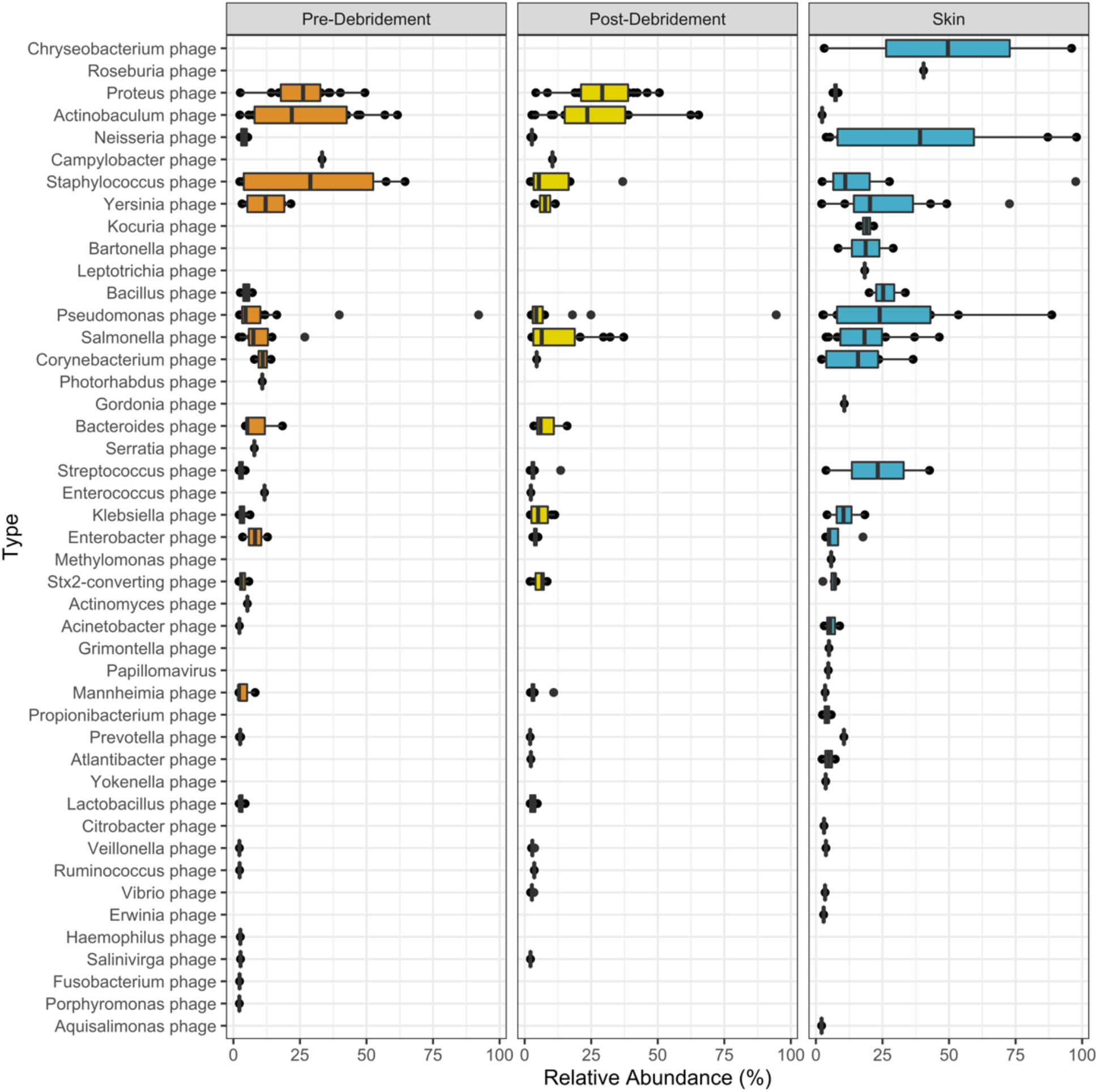
Taxonomic composition of viruses identified in wound (pre-debridement and post-debridement) and skin samples. Relative abundance is given for each taxon > 2% at the ‘type’ level (presumed host designation). Center line, median; box limits, upper and lower quartiles; whiskers, 1.5x interquartile range; points, outliers; *n* = 20 patients.

Within the wound samples, the most prominent presumed phage hosts were *Proteus, Actinobaculum, Staphylococcus, Campylobacter, Yersinia, Pseudomonas* and *Salmonella* (Figure 1). For viruses that could be identified at the species level, wounds had high abundances of *Proteus* phage VB PmiS, an unnamed *Actinobaculum* phage, *Pseudomonas* virus phiCTX, and *Staphylococcus* phages StauST398-5 and Sextaec (Figure S6). The top viral types shared between skin and wounds are presumed to infect *Staphylococcus, Yersinia, Pseudomonas* and *Salmonella*.

### Viral diversity is greater in wounds compared to skin

The virome exhibits significantly higher intra-sample taxonomic richness and evenness in wounds than skin, as measured by alpha diversity metrics with unclassified taxa included (Figure 2a). In terms of richness, wounds had an average Chao1 index of 996 ± 426 while skin had an average of 101 ± 271. Accounting for abundance and evenness, wounds had an average Shannon index of 4.70 ± 0.72 while skin had an average of 1.95 ± 1.23. The differences in richness and evenness can be qualitatively visualized in a relative abundance heatmap of the top 300 taxa (Figure 2b). The findings contrast with diversity of the bacterial fraction of skin and wound microbiomes, in which bacterial diversity is consistently higher in normal skin compared to wounds ([26]; Figure 2c). To eliminate the possibility that increased richness was due to greater sampling, we subsampled each sample to an equal depth and performed the same diversity analysis. The trends in phage diversity remained the same (higher diversity in wounds compared to skin) (Figure S7), indicating that the differences were not driven by sample size.

**Figure 2.**
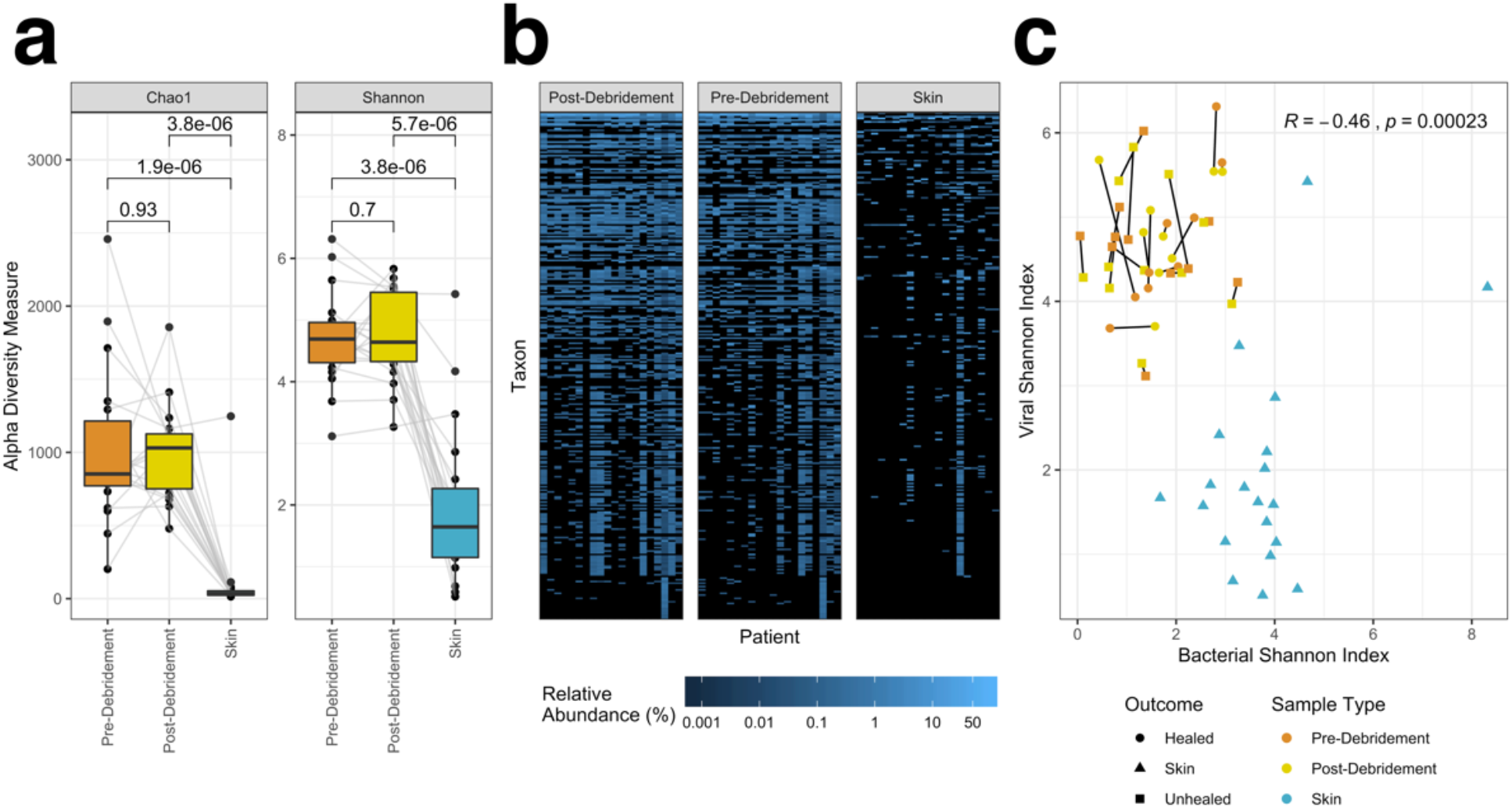
Alpha diversity of skin and wound viromes. (a) Boxplots of Chao1 and Shannon indices in wound samples (pre- and post-debridement) and skin samples, with each patient’s samples connected by grey lines. Center line, median; box limits, upper and lower quartiles; whiskers, 1.5x interquartile range; points, outliers; *n* = 20 patients. Averages were compared with paired, two-sided Wilcoxon signed-rank tests, resulting in the p-values shown. (b) Heatmap of relative abundance of the top 300 taxa in each patient’s pre-/post-debridement and skin samples. (c) Correlation between viral and bacterial Shannon indices, with pre- and post-debridement samples from the same wound connected by a black line, sample type indicated by color, and outcome indicated by shape. Pearson’s correlation coefficient R and p-value are shown, calculated using all samples.

### Skin and wound viromes are taxonomically distinct

Diversity between samples (beta diversity) was measured using Bray-Curtis distance and visualized by principal coordinates analysis. Unclassified taxa were included. Skin and wound samples partitioned well from each other (Figure 3a), indicating that they harbor distinct viromes. As observed previously for the bacterial fraction, pre- and post-debridement wound samples were more similar to each other than to the corresponding skin sample (Figure 3b).

**Figure 3.**
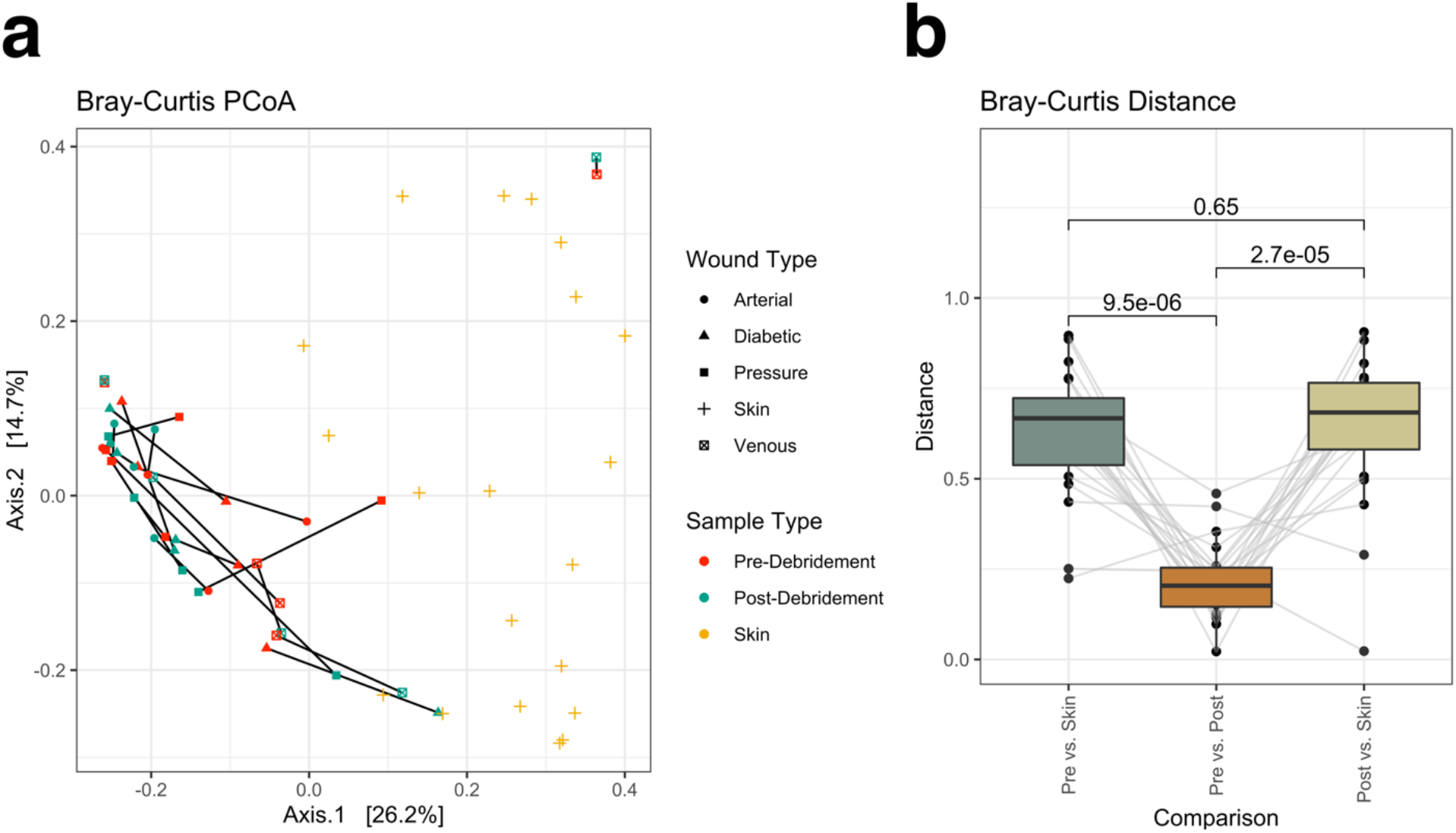
Beta diversity as measured by Bray-Curtis dissimilarity. Taxa present in > 2 samples with > 0.5% relative abundance (including unclassified taxa) were retained for analysis. Ordination of the Bray-Curtis dissimilarity matrix using principal coordinates analysis (a) illustrates distinct compositions for wound and skin samples. Within-patient dissimilarity between pre-debridement, post-debridement, and skin samples with averages were compared by two-sided Wilcoxon signed-rank tests (p-values shown) and data from each patient are connected by grey lines (b). Center line, median; box limits, upper and lower quartiles; whiskers, 1.5x interquartile range; points, outliers; *n* = 20 patients.

### Specific viral species and hosts associated with skin and wound samples

Species associations to skin and wound sample types were also explored by differential abundance analysis using DESeq2. Although unique phage species were associated with each sample type, the hosts they targeted were largely shared, including *Yersinia spp*., *Neisseria spp*., *Pseudomonas spp*., *Streptococcus spp*., *Salmonella spp*., and *Staphylococcus spp*. (Figure 4). Nevertheless, skin and wounds differed in some of the host species targeted. Skin was associated with one *Staphylococcus haemolyticus* phage, one *Staphylococcus aureus* phage, and two general *Pseudomonas* phages, while wounds had many associations to *Staphylococcus aureus* and *Pseudomonas formosensis* phage species.

**Figure 4.**
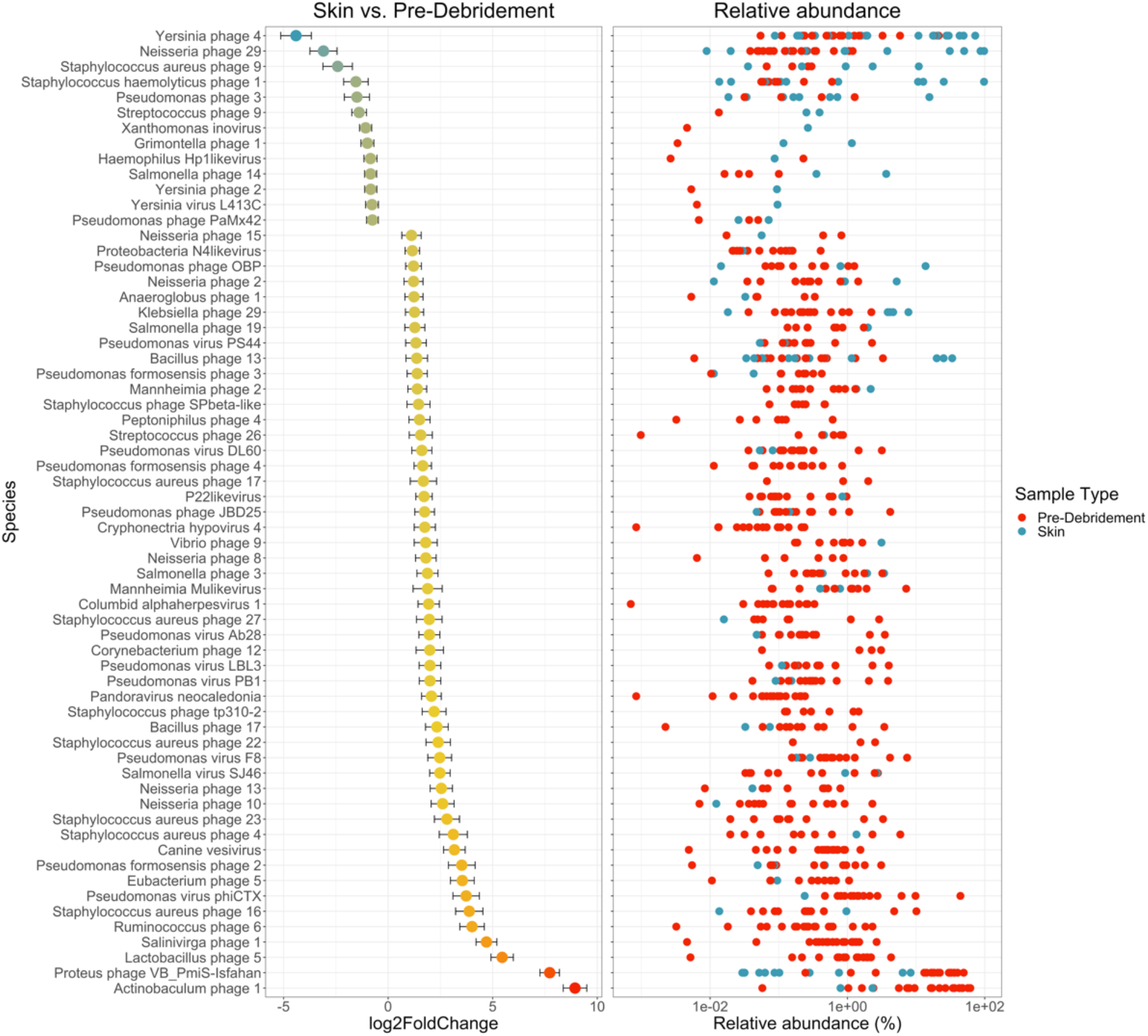
Differential abundance analysis of skin and chronic wound viromes with DESeq2. Associated species are represented by their log2FoldChange from the normalized, geometric mean calculated across all wound samples, contrasting skin (negative change) to wounds (positive change). Error bars represent the log-fold standard error; *n* = 20 patients. Only species with adjusted p-value < 0.05 are shown. Relative abundance of each associated species, in each sample, are shown on the right; wound samples (pre-debridement) are red, skin samples are blue.

### Viral species associated with healing outcomes

To identify specific taxonomic associations to covariates, differential abundance analysis was performed with DESeq2 (Figure 5). Wound were classified according to whether they healed within 6 months after sampling. Within wound samples, associations to healing outcomes are of primary interest. After filtering the results to retain associations with adjusted *p*-values < 0.01, both healed and unhealed wounds were found to be associated with specific *Staphylococcus* phage. Healed wounds were also associated with many *Pseudomonas, Campylobacter* and *Bacteroides* phage, while unhealed wounds were associated with *Enterococcus, Enterobacter, Veillonella* and *Streptococcus* phage. Host association was known for these phages, but most did not have species designations approved by the International Committee on Taxonomy of Viruses (ICTV).

**Figure 5.**
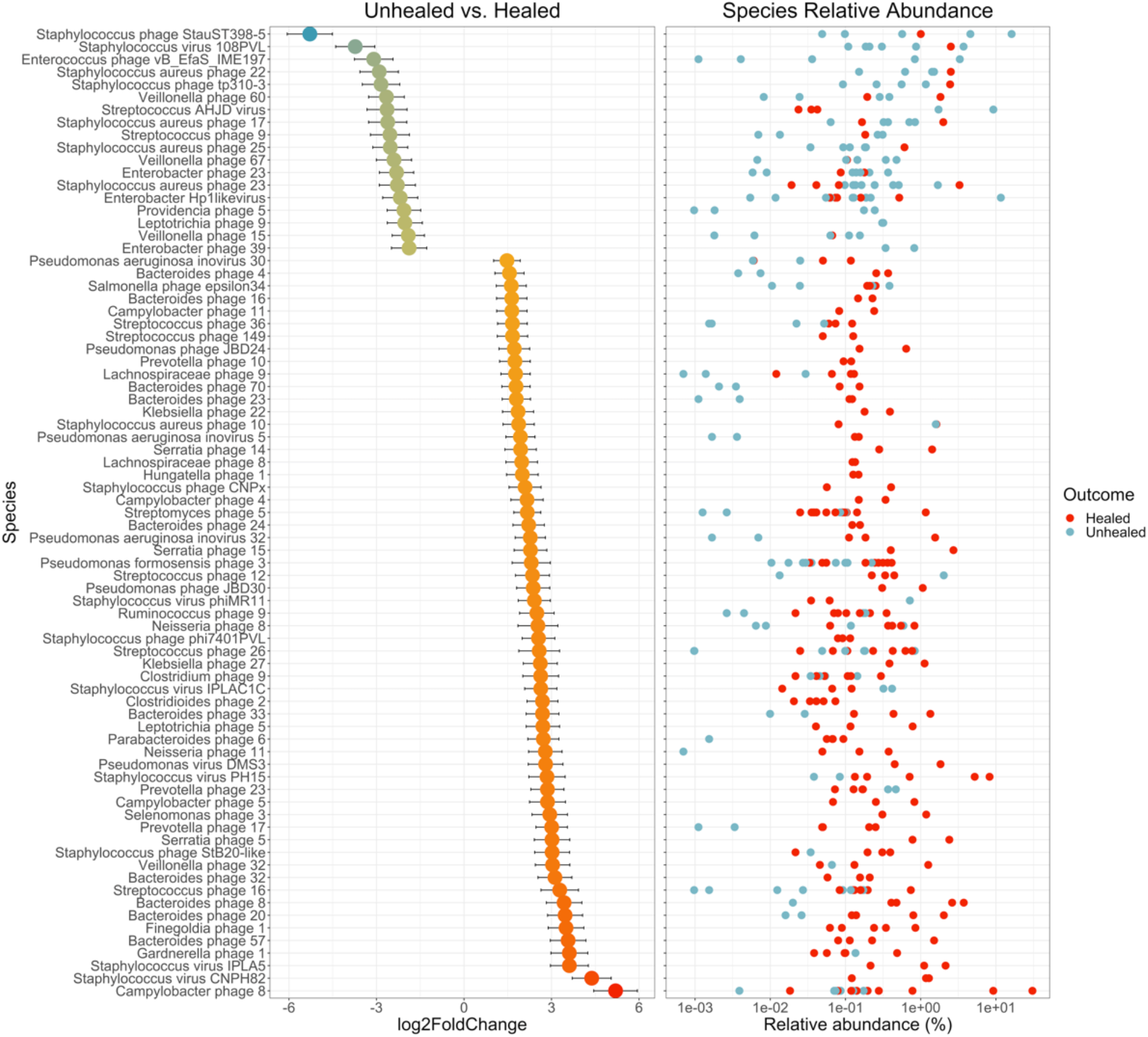
Differential abundance analysis of healed and unhealed wound viromes with DESeq2. Associated species are represented by their log2FoldChange from the normalized, geometric mean calculated across all wound samples, contrasting unhealed (negative change) to healed wounds (positive change). Error bars represent the log-fold standard error; *n* = 20 patients. Only species with adjusted p-value < 0.01 are shown. Relative abundance of each associated species, in each sample, are shown on the right; healed wound samples are red, unhealed wound samples are blue. Numeric species designations given were assigned by this study.

## Discussion

Using deep sequencing, we characterized the skin and chronic wound viromes of 20 patients presenting at a wound care clinic. Samples were processed using a virus-like particle (VLP) enrichment protocol to capture actively replicating viruses, reduce host contamination and increase viral sequencing depth. Here we report the taxonomic composition of the viromes, associated ecological diversity measures, and specific taxa associated with healing outcomes and sample types.

Host contamination was assessed by classifying reads against the full NCBI RefSeq database, indicating a high abundance of human and bacterial DNA. It should be noted that this assessment underestimates true viral read abundances, which may be aligned to CRISPR spacers and prophages in bacterial reference genomes [45, 46]. Additionally, since NCBI’s viral reference database is incomplete, unclassified reads may have viral origin. Despite the implementation of VLP-enrichment procedures and attempted depletion of host DNA, the prevalence of human and bacterial contamination underscores the need for extensive host DNA degradation during sample processing.

Viral detection and classification were performed with unassembled reads in a *k*-mer-based, reference-dependent manner. A viral metagenome database was utilized to capture relatively new or poorly annotated viruses. Nevertheless, approximately half of viral taxa detected had no known host or species annotation, similar to previous work in the field [37, 47]. These results are consistent with a large, unclassified fraction of viral material, emphasizing the importance of read assembly, protein homology searches, and other methods for characterizing the virome [45, 48-52]. Regardless, of the annotated viral taxa, most targeted abundant skin and wound bacteria such as *Proteus spp*., *Actinobacteria*., *Pseudomonas spp*., *Staphylococcus spp*., *Corynebacterium spp*., *Streptococcus spp*. and mixed Proteobacteria. Wound samples tended to contain pathogen-targeting phages, while skin samples were associated with phages targeting commensals, consistent with the bacterial taxa present in the respective sample types previously detected using 16S rRNA sequencing [26].

Diversity analysis, including the unannotated viral taxa, showed that wound viromes were significantly more diverse than skin viromes in both taxonomic richness and evenness. Interestingly, viral diversity was negatively correlated with bacterial diversity. This correlation did not appear to be an artifact of low viral sampling depth of skin, since subsampling to the same depth gave similar results (Figure S7). The findings suggest that the wound environment, while resulting in low bacterial diversity, may be hospitable to proliferation of diverse phages. Wound treatment may also encourage phage proliferation, as lysogenic phage may switch to the lytic life cycle in response to antibiotics [53], reactive oxygen species [54], DNA damage signaled by SOS responses [55], and various stress responses to changes in the environment, like pH [56, 57]. Skin and wound viromes were also non-linearly partitioned by beta diversity ordination, illustrating that their compositions were distinct.

To characterize possible taxonomic associations to wound healing status, wounds were classified as healed (8 wounds) or unhealed (12 wounds), based on whether the wound had healed within 6 months after sampling. Differences between the viromes of healed and unhealed wounds were characterized by differential abundance analysis. Several significantly associated taxa displayed unique functional properties that could influence healing outcomes (Table 1). Both healed and unhealed wounds were largely associated with temperate phage in the family Siphoviridae, including *Staphylococcus* phages carrying Panton-Valentine leukocidin genes [58, 59]. Their presence in wounds may indicate a shift to the lytic cycle in the wound environment. Furthermore, as temperate phages, Siphoviridae may exert influence over their hosts’ function through prophage integration and lysogenic conversion [31, 60]. Of the lytic phages, healed wounds were associated with two *Staphylococcus* phage species, and unhealed wounds were associated with *Streptococcus* and putative *Enterobacter* phage species. Phage species associated with healed wounds may have profound impacts on host function, including reduction or inhibition of biofilm formation [61, 62], motility inhibition equivalent to a pilus knockout [61], CRISPR resistance or inhibition [62, 63], and anti-biofilm activity via capsid-displayed pectin lyase-like domains [64]. Biofilms are a leading impediment to wound healing, and exploitation of phage or their proteins as anti-biofilm agents is a very active area of research [34, 65-67]. Suppression of the CRISPR system might confer an advantage to the phage in evading degradation by the host. Recent work suggests that anti-CRISPR systems are dependent on multiplicity of infection, requiring several phage to be expressing the gene simultaneously in a rare case of inter-phage cooperation and altruism [68]. The association of phages bearing such traits with healing of chronic wounds may warrant further investigation.

**Table 1.** Traits of viruses associated with healed and unhealed wounds.

| Association | Species | Known hosts | Lifecycle | Notable traits | Reference |
| --- | --- | --- | --- | --- | --- |
| Healed | *Pseudomonas* phage JBD24 | *Pseudomonas aeruginosa* | Temperate | inhibits motility (equivalent to pilus knockout); reduced biofilm formation | [61] |
| Healed | *Pseudomonas* phage JBD30 | *Pseudomonas aeruginosa* | Temperate | inhibits CRISPR systems | [63] |
| Healed | *Pseudomonas* phage DMS3 | *Pseudomonas aeruginosa* | Temperate | inhibits biofilm formation; exhibits CRISPR resistance | [62] |
| Healed | *Salmonella* phage epsilon34 | *Salmonella enterica* subsp. *enterica* serovar | Temperate | P22/Lambdaoid phage (potential contaminant); alters *Salmonella* spp. serotype | [69] |
| Healed | *Staphylococcus* phage CNP<sub>x</sub> | *Staphylococcus epidermidis* | Temperate |  | [70] |
| Healed | *Staphylococcus* phage phi7401PVL | *Staphylococcus aureus* | Temperate | carries pore-forming toxin PVL | [58] |
| Healed | *Staphylococcus* phage StB20-like | *Staphylococcus epidermidis*; *Staphylococcus hominis* | Temperate |  | [71] |
| Healed | *Staphylococcus* phage CNPH82 | *Staphylococcus epidermidis* | Temperate |  | [72] |
| Healed | *Staphylococcus* phage IPLA5 | *Staphylococcus epidermidis* | Lytic | pectin lyase-like domains; anti-biofilm activity | [64] |
| Healed | *Staphylococcus* phage IPLAC1C | *Staphylococcus* ssp. | Lytic |  | [73] |
| Healed | *Staphylococcus* phage PH15 | *Staphylococcus epidermidis* | Temperate |  | [72] |
| Healed | *Staphylococcus* phage phiMR11 | *Staphylococcus aureus* | Temperate |  | [74] |
| Unhealed | *Enterobacter* Hp1likevirus | *Enterobacter* | Lytic |  | [75] |
| Unhealed | *Enterococcus* phage vB_EfaS_IME197 | *Enterococcus faecalis* | Temperate |  | [76] |
| Unhealed | *Staphylococcus* phage tp310-3 | *Staphylococcus aureus* | Temperate |  | [77] |
| Unhealed | *Staphylococcus* phage108PVL | *Staphylococcus aureus* | Temperate | carries pore-forming toxin PVL | [59] |
| Unhealed | *Staphylococcus* phage StauST398-5 | *Staphylococcus aureus* | Temperate |  | [78] |
| Unhealed | *Streptococcus* phage AHJD | Group C *Streptococci* | Lytic |  | [79] |

Both healed and unhealed wounds were associated with phage known to transduce the pore-forming toxin Panton-Valentine leukocidin (PVL), which may increase pathogenicity of their hosts by evading immune response and lysing leukocytes, though the specific role of leukocidins in wound pathogenesis is still unclear [80]. The Proteus phage vB PmiS-TH was also prominent among wound samples. This Siphoviridae phage has been found to be lytic against *P. mirabilis*, a known wound pathogen, and may be a common wound commensal phage [81]. Future studies would be needed to characterize any such possible functional associations.

Several considerations limit the analysis presented here. The protocols used here are appropriate for non-enveloped dsDNA viral particles, possibly including replicative intermediates of ssDNA viruses. Other viruses, including prophages and RNA viruses, would require further investigation. We determined viral abundance and taxonomy using a k-mer-based approach at the nucleotide sequence level with unassembled reads. Viral detection, taxonomic classification, and associations were therefore limited by existing reference databases. The NCBI Viral RefSeq database is relatively small, but curated and annotated, while IMG/VR is large but less annotated. In particular, temperate bacteriophage may excise portions of the host genome when entering the lytic lifecycle [31, 57]; if such bacterial sequences were present in the IMG/VR database, bacterial contaminants in the sample data may have appeared as false-positive viral hits. In addition to this, the sequences contained a substantial amount of human and bacterial DNA despite the use of viral enrichment protocols, and *in silico* decontamination was needed. Contamination is a common issue for viral metagenomics, especially when working with low-biomass clinical samples [82, 83]. Contamination was more prevalent for skin samples, as expected given the lower DNA yields. Additional measures, such as more robust nuclease treatment and improved *in silico* decontamination methods, could improve sequence quality in future work. Finally, due to the small cohort size, only associations of relatively large effect could be detected, and the findings of this study should ideally be validated with larger cohorts.

## Conclusion

Chronic wounds are frequently colonized and infected by polymicrobial communities, impeding wound healing. Previous work has established that the bacterial fraction of these communities exhibits high interpersonal variance, and community structure and function may be associated with healing outcomes. Yet the forces that drive compositional and functional dynamics of wound microbiomes have yet to be elucidated. We sought to better understand the role of a potentially important contributing factor, the virome. This study presents the first characterization of the chronic wound virome, utilizing a virus-like particle enrichment protocol and shotgun metagenomics to survey the wounds of 20 patients presenting at an outpatient wound care clinic. Despite heavy host contamination, we describe viral taxonomic composition, diversity, associations to covariates, and virus-host correlations.

While no causative or conclusive claims can be made regarding the virome’s role in wound pathology, the rich inter- and intra-personal taxonomic diversity and associations to covariates suggest that the virome is a prominent component of the greater microbiome and merits thorough investigation in the future. To achieve more sensitive viral detection and accurate taxonomic classification, future studies would benefit from shotgun sequencing both the bacterial and viral fractions of the microbiome, and assembling the resulting reads into contiguous sequences, which will facilitate protein homology searches and within-sample CRISPR spacer and prophage alignments. Furthermore, time series data will be imperative for elucidating the multitude of complex, dynamic bacteria-bacteriophage interactions. Such work will contribute to the greater understanding of how the wound microbiome as a whole is related to wound pathology, and ultimately, how it may be leveraged to achieve more positive healing outcomes.

## Methods

### Ethics Statement

Clinical sample collection was performed at Ridley-Tree Center for Wound Management at Goleta Valley Cottage Hospital in accordance with protocols approved by the Cottage Health Institutional Review Board (Study Protocol 17-48u) and UCSB’s Human Subjects Committee and Institutional Review Board (Study Protocol 4-18-0190). A cohort of 20 wound care patients were recruited over the course of a week and a half, and samples were collected after obtaining informed, written consent from the patient.

### Clinical sample collection

Samples were collected as previously described [26]. Four clinically classified chronic wound types were sampled (diabetic ulcers, venous wounds, arterial wounds, and pressure ulcers), with five patients per wound type. Exclusion criteria were: patients under the age of 18, in the intensive care unit, or presenting with an unrelated non-wound infection. All patients underwent non-conservative sharp debridement until bleeding was observed. However, the extent and depth of debridement, as well as the type of instrument (curette, scalpel, scissors, or tissue nipper), was not standardized and was determined by the treating physician (see [26]). Sterile Copan FLOQSwabs 520C were pre-wetted with sterile PBS prior to all sample collections. During a single patient visit, wound swabs were collected pre-debridement and 1-2 minutes post-debridement, and a healthy skin swab was collected from the contralateral limb. Wound samples were collected from the area of debridement. All skin and wound samples were collected by employing Levine’s technique; gentle pressure was applied as the swab was wiped and rolled across a ∼ 1cm^2^ area of healthy granulation tissue for approximately 30 seconds. Clinical swabs were placed back into the dry, sterile collection tube and stored at 4°C for no more than four hours before being processed. Negative control samples from the wound center were collected by exposing swabs to air in the collection room for the same duration as wound and skin swab collection. Processing control samples were obtained by exposing swabs to air and reagents in the processing lab analogously to clinical samples.

### Sample processing and DNA extraction

Samples were processed as described [39]. Briefly, swab tips were inserted into 1.5 mL microcentrifuge tubes and snapped at the 30 mm break-point. 500 µL of sterile 1× TE was added to the tube, and the tube was vortexed for 2 minutes at maximum. Speed on a multitube vortex adapter to resuspend the sample. Samples were then centrifuged at 16,000 × g for 2 minutes to pellet cells. 250 µL of supernatant was transferred to a 2mL microcentrifuge tube for immediate VLP precipitation. The remaining 250 µL of supernatant, pelleted cells, and swab tip were kept in the original tube and stored at −20°C before proceeding to whole-microbiome DNA extraction.

### Isolation of DNA from virus-like particles

VLP purification and DNA extraction was conducted as described [39]. Briefly, free DNA in the VLP fraction was digested with DNase I (5 units, NEB) at 37 °C for 30 minutes; DNase I was inactivated by incubation at 75 °C for 10 minutes. VLPs were precipitated, pelleted, washed with ice cold 70% ethanol, and re-pelleted. Pellets were dried for 1 hour at room temperature in a vacufuge before being resuspended in sterile 1X TE (pH 8.0). Viral capsids were disrupted and digested with 10% SDS and proteinase K, incubated at 55°C for 1 hour. VLPs were further disrupted with 5M NaCl and CTAB-NaCl, followed by incubation at 65°C for 10 minutes. The sample was then transferred to a phase lock gel tube (5PRIME PLG Light) and mixed with 250 μL of 25:24:1 phenol:chloroform:isoamyl alcohol by inversion. Phases were separated by centrifugation at 1500 × g for 5 minutes. In the same tube, 24:1 chloroform: isoamyl alcohol extraction was performed twice and centrifuged as described above, and the 250μL aqueous phase was transferred to a 2mL microfuge tube. DNA was purified by ethanol precipitation. Pellets containing DNA were washed with 500μL ice cold 70% ethanol, and re-pelleted by centrifugation, then dried for 1 hour at room temperature in a vacufuge and resuspended in 20 μL 1X TE (pH 8.0).

### Library preparation and sequencing of VLP DNA

DNA from VLP-enriched samples was quantified using the Qubit dsDNA HS kit. Two library preparation methods were utilized depending on DNA concentration. Both methods are based on the Nextera XT kit with Nextera XT V2 set A indices. Samples with DNA concentrations > 0.2 ng/μL (43/66 samples) were diluted and normalized to 0.2 ng/μL and prepared for shotgun sequencing as described by the manufacturer. Samples with DNA concentrations < 0.2 ng/μL (23/66 samples) were prepared for shotgun sequencing using a ‘tagmentation’ reaction modified and optimized for low-input samples, as described in [84]. All indexed samples were quantified with the Qubit dsDNA HS kit, normalized, and pooled. A final, double size-selection step was performed using AMPureXP beads. Final library quality control was done using Agilent TapeStation dsDNA 5000 bp and 1000 bp kits. Final libraries were sequenced on an Illumina HiSeq 4000 with PE150 V3 chemistry, using two lanes, at the UC Davis DNA Technologies Core.

### Viral read pre-processing

Initial quality analysis was performed with FastQC. Read pre-processing was performed by quality trimming, adapter trimming, quality filtering, and length filtering with trimmomatic using Nextera XT adapter sequences and ‘palindrome’ mode for adapter trimming; all other settings were defaults [85]. Trimmed, paired reads were joined with PANDASeq with default parameters [86]. Trimmed singletons and joined pairs were concatenated together into the final pre-processed read set for each sample.

### Taxonomic read classification and abundance estimation

Overall taxonomic read classification (eukaryotic, bacterial, archaeal, and viral) was performed on pre-processed reads at the nucleotide level against the full NCBI RefSeq database with Kraken2 [40, 87]. For each sample, species abundances were estimated using the Bracken package with an ideal read length of 150bp [41]. To better characterize the viral read content, pre-processed reads were first classified against NCBI’s Viral RefSeq database with Kraken2 [40, 43]. The remaining, unclassified reads were re-classified against the full IMG/VR database (IMG VR 2018-07-01 4) with Kraken2 [40, 42]. For each sample in each viral classification method, species/taxon abundances were estimated with Bracken using an ideal read length of 150bp [41]. Abundance reports for each sample in each viral classification method were combined into a single count table. Viral and host taxonomy were abstracted from NCBI and IMG/VR, and manually curated to standardize viral species and ‘type’ level strings. For NCBI taxa, host association was inferred from the viral species designation, while IMG/VR host assignments were determined by a combination of viral species designation, alignment to CRISPR spacers and prophages, and deposited metadata. For taxa without a species designation, the ‘type’ designation with a numerical ID was used. Additional information, like putative phage morphology, was inferred from IMG/VR viral cluster metadata. After curation, NCBI and IMG/VR taxonomy tables were concatenated to create a single taxonomy table for downstream analyses.

### Viral community composition and differential abundance analyses

The combined Bracken count table, taxonomy table, and a metadata mapping file were imported to RStudio and built into a Phyloseq object for community composition analyses [88]. Contaminants were detected and identified using Decontam, with four negative control samples and a threshold of 0.2 [44]. Additional decontamination was performed by filtering viral species, strains and types known to be used in adjacent laboratory space; prominent contaminants are shown in Figure S3. A large proportion of unannotated taxa remained after decontamination; unless otherwise stated, all analyses were performed with this fraction removed. Stacked taxonomic boxplots were generated with phyloseq after agglomeration taxa at the species or host/type level. Alpha diversity was calculated with phyloseq [88], plotted with ggplot2 [89], and stats calculated with ggpubr. Beta diversity was calculated and ordinated with phyloseq; additional boxplots were made with ggplot2. All additional analyses and visualizations of community composition were performed using a combination of Phyloseq, dplyr, ggplot2, and ggpubr. Differential abundance analyses were performed with DESeq2 using non-parametric fitting, the Wald test for significance, and the Benjamini-Hochberg correction for multiple hypothesis testing [90]. Results were visualized with ggplot2, with error bars representing the log-fold standard error.

### 16S rRNA library preparation, sequencing and bioinformatics

16S rRNA library preparation, sequencing, and bioinformatics were performed as previously described [26, 39]. Briefly, the whole-microbiome fraction was extracted by enzymatic digestion with high-activity lysozyme and proteinase K, followed by incubation with chemical lysis buffer and mechanical lysis by bead beating. Extracted DNA was purified using the PureLink Genomic DNA kit, following the manufacturer’s instructions. Sequencing libraries were prepared using 2-step PCR, targeting the V1-V3 loops of the 16S gene, and libraries were sequenced on an Illumina MiSeq with a PE300 kit. Reads were processed with QIIME using the open OTU picking pipeline [91], and taxonomy was assigned against the SILVA128 database [92]. The resulting BIOM table was imported to RStudio, along with a mapping file, and built into a phyloseq object for downstream analyses [88].

## Data Availability

All data produced in the present study are available upon reasonable request to the authors

## Acknowledgements

Funding for this project was provided by the NIH New Innovator Program [Grant Number DP2GM123457] and the Camille Dreyfus Teacher-Scholar Program. We thank Y. Shen for computational insights.

## Author contributions

SV, JMD and IAC designed the study. SV conducted experiments and performed computational analysis. JMD collected patient samples. IAC provided research direction. SV and IAC wrote the manuscript with input with all authors.

