## Supplementary Information File for "The chronic wound virome: phage diversity and associations with wounds and healing outcomes"

### Supplementary Data

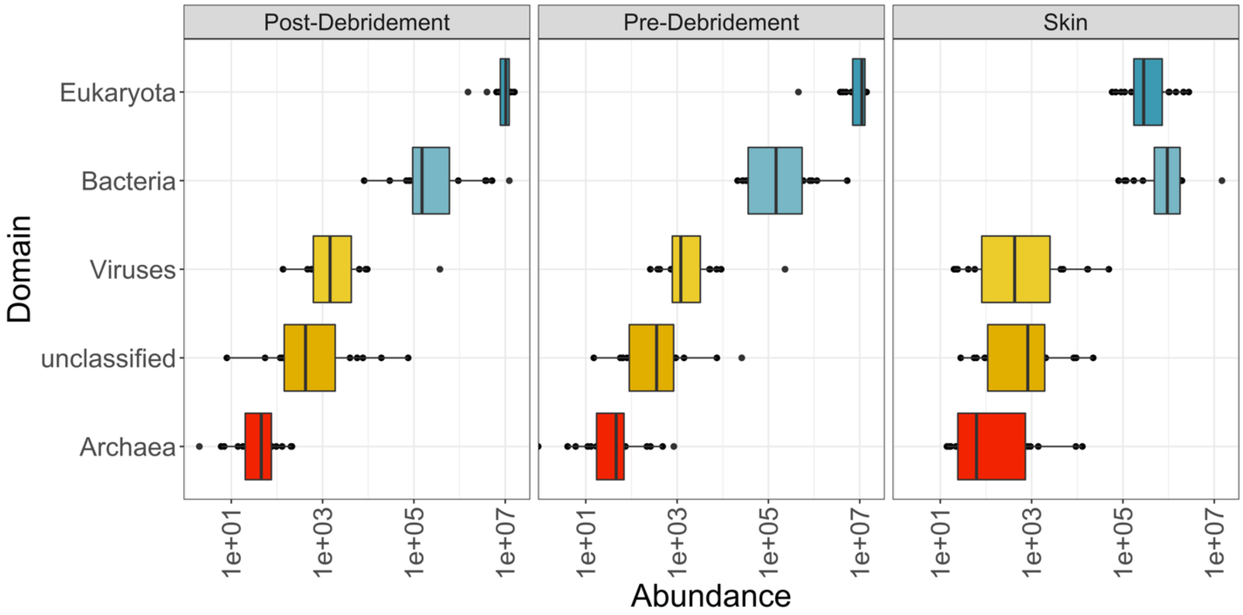


Figure S1. Domain-level taxonomic composition of metagenomic samples. Read abundances in each domain for each sample type, as classified by Kraken2 against the full NCBI RefSeq database. Samples contained 6,794,663 ± 2,588,627 eukaryotic and 1,205,730 ± 2,588,627 bacterial reads, on average across all samples. 13,278 ± 55,825 reads were classified as viral, on average. An additional 3,660 ± 10,720 reads remained unclassified, on average. Center line, median; box limits, upper and lower quartiles; whiskers, 1.5x interquartile range; points, outliers; *n* = 20 patients.

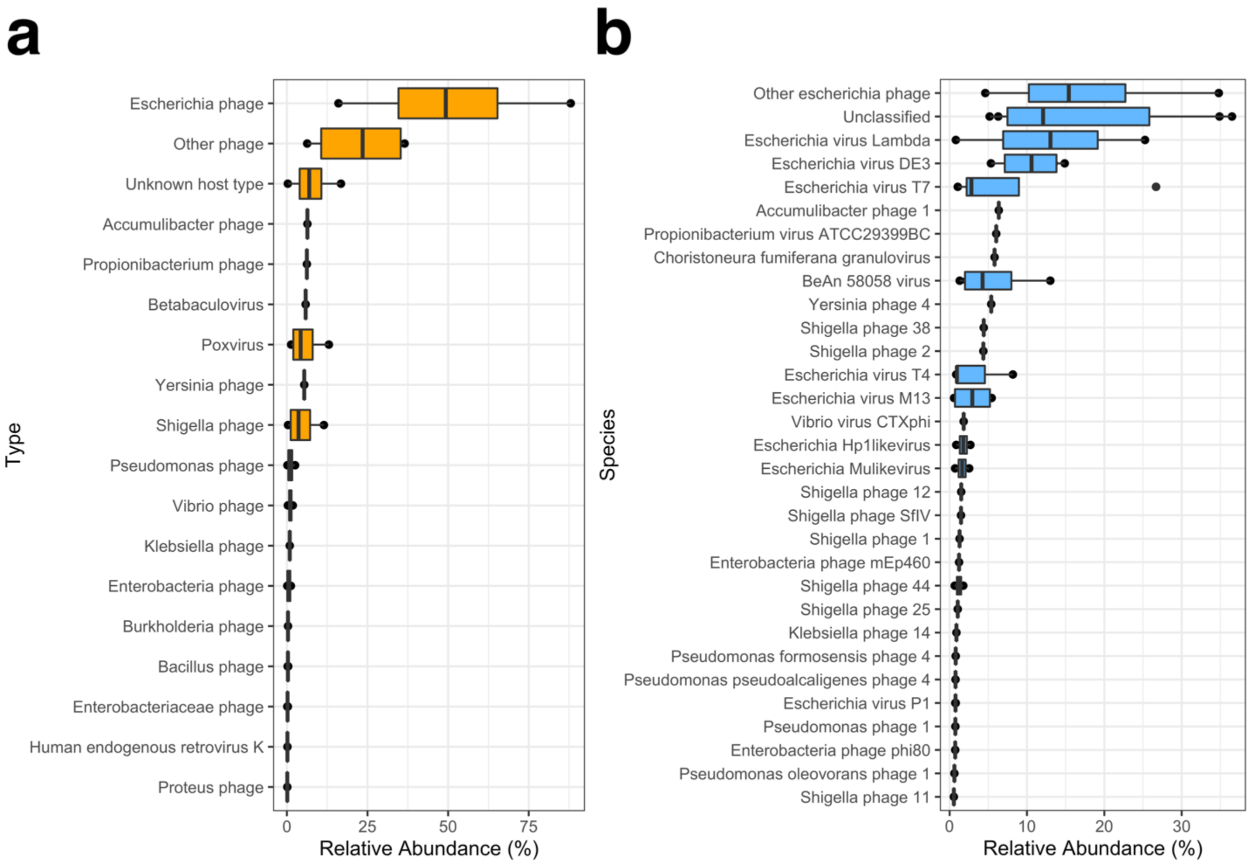


Figure S2. Negative control composition boxplots. Relative abundance of each taxon in each of the four negative control samples, prior to decontamination; only taxa with > 0.1% are shown at the type level (a) and > 0.5% at the species level (b). Center line, median; box limits, upper and lower quartiles; whiskers, 1.5x interquartile range; points, outliers; *n* = 20 patients.

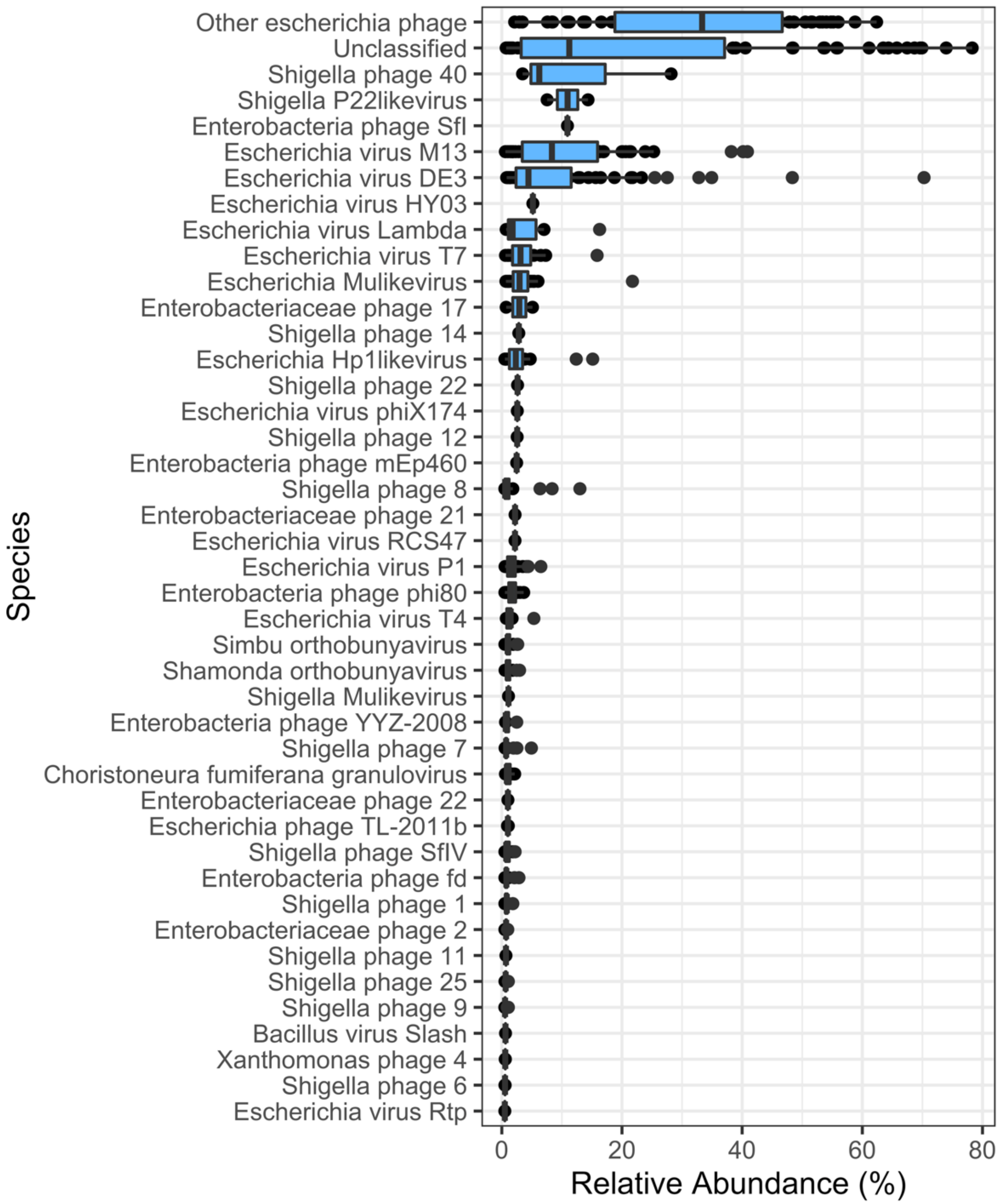


Figure S3: Contaminant relative abundance in all skin and wound samples prior to decontamination. Taxa displayed were designated as contaminants with relative abundance > 0.5% in > 1 skin or wound sample prior to decontamination. Center line, median; box limits, upper and lower quartiles; whiskers, 1.5x interquartile range; points, outliers; *n* = 20 patients.

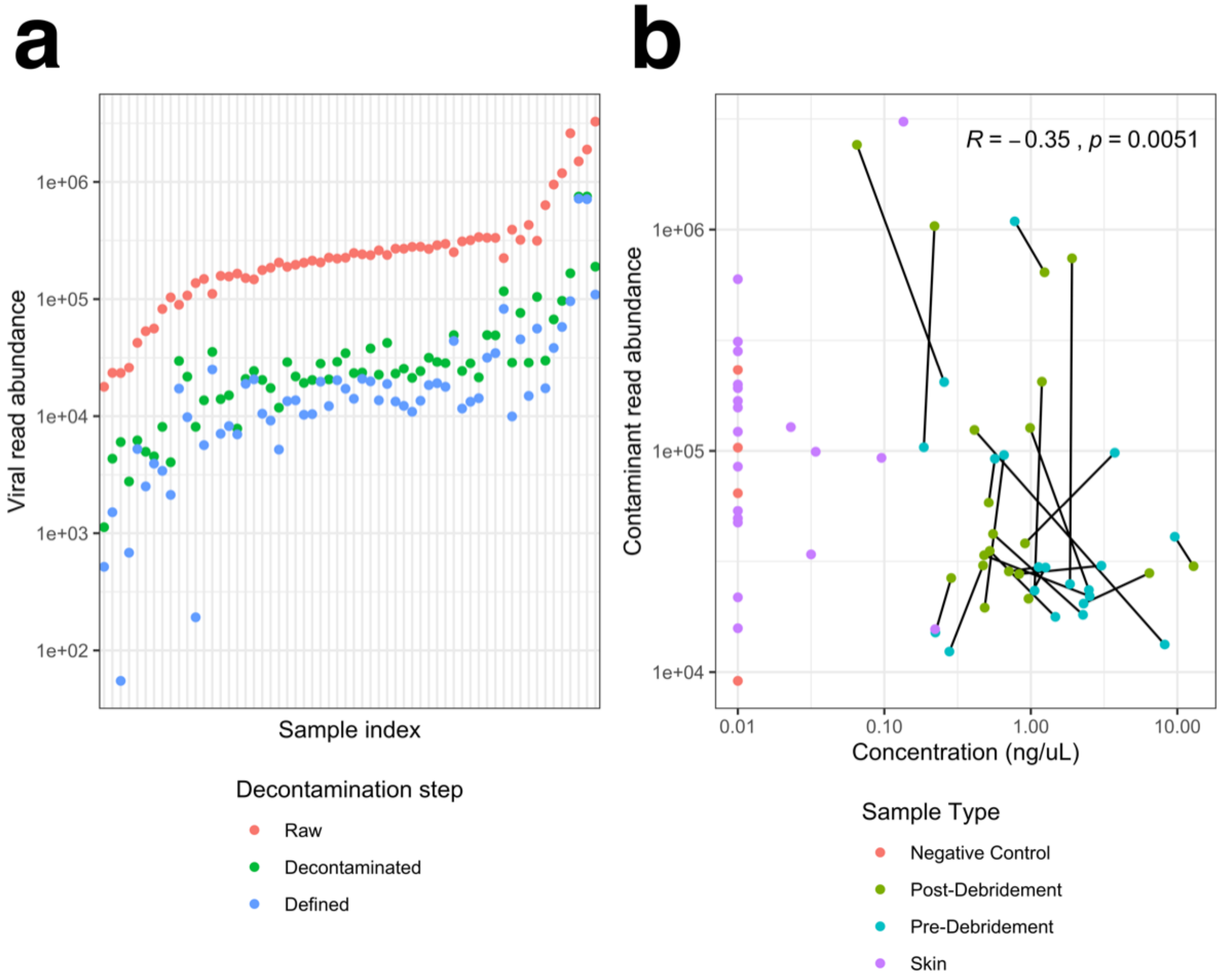


Figure S4. Decontamination summary. Read abundances for each sample, at each step of decontamination (a). Only experimental samples are shown, not negative or positive controls. Raw is the number of intial reads, decontaminted is the number of reads after contaminants are removed, and defined is the number of reads with a taxonomic or host classification. (b) Contaminant read abundance as a function of extracted DNA concentration, with points colored by sample type, and wound samples from the same patient connected by black lines. Overall Spearman correlation coefficient R and p-value are shown.

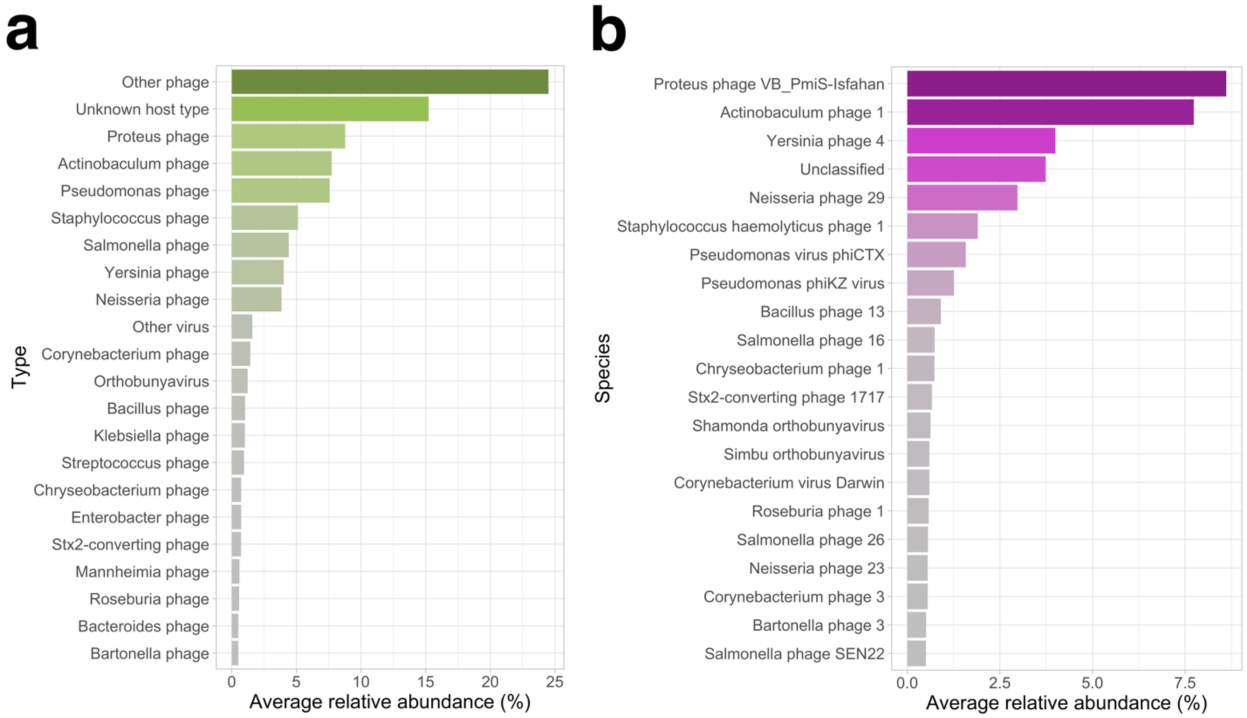


Figure S5. Overall taxonomic composition of skin and wound viromes with unclassified taxa included. Relative abundance of each taxon was averaged across all samples; those with > 0.5% average relative abundance are shown here at the type level (a) and species level (b).

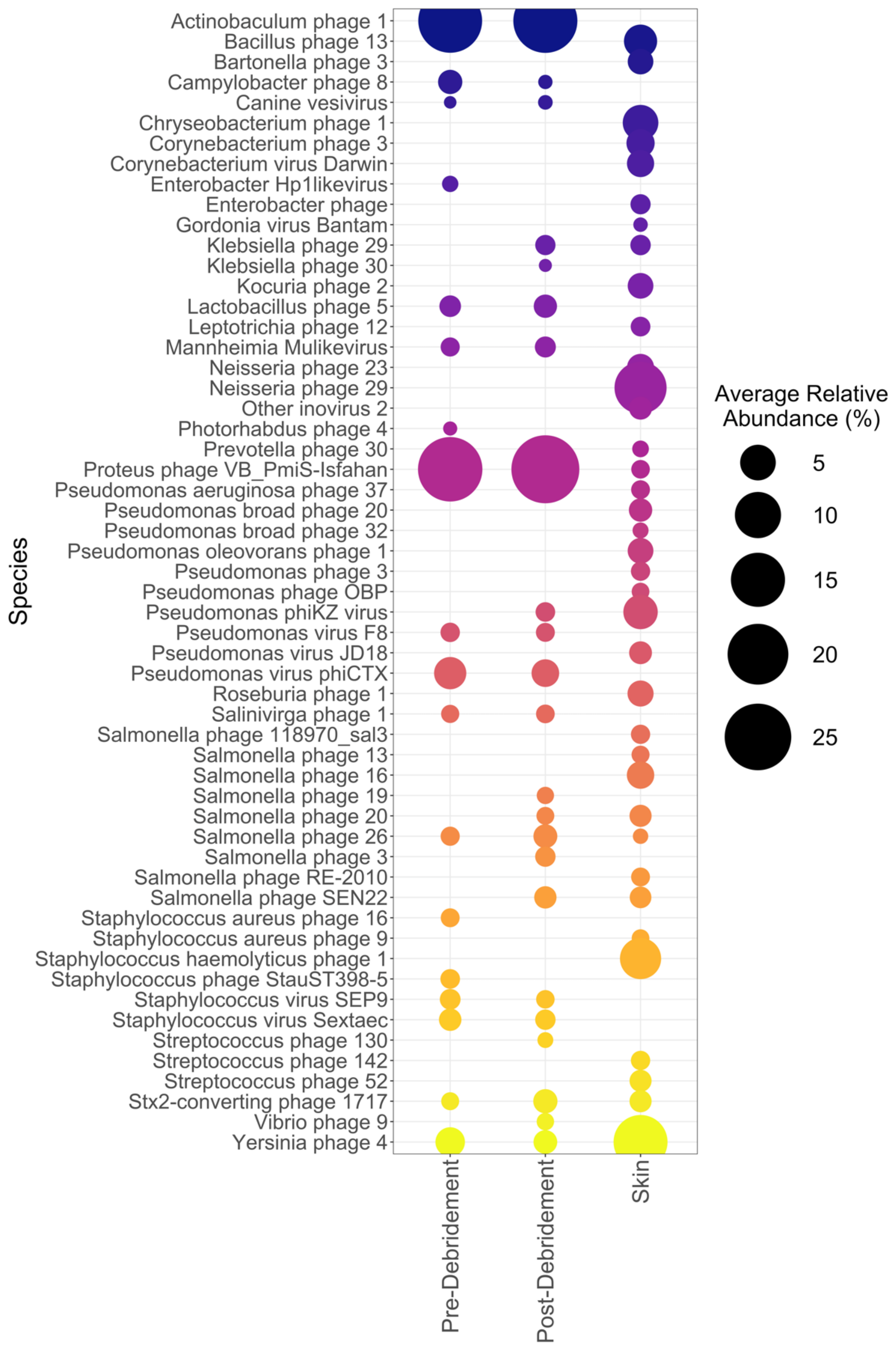


Figure S6. Average relative abundance of viral species by sample type. Relative abundances were averaged within pre-debridement, post-debridement, and skin sample types; taxa with average relative abundance > 0.5% are shown.

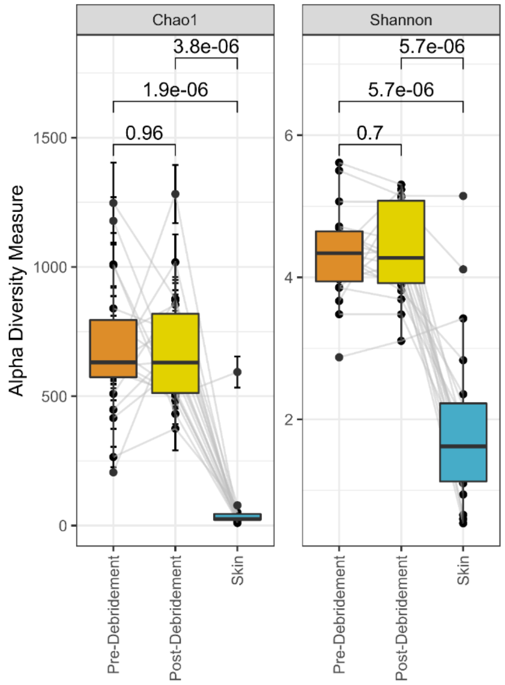


Figure S7: Subsampled alpha diversity analysis. Abundance values were subsampled at an even depth equal to that of the sample with the minimum total abundance. Center line, median; box limits, upper and lower quartiles; whiskers, 1.5x interquartile range; points, outliers; $n$ = 20 patients.
